# Next-generation sequencing reveals the presence of a rich bacterial microbiome in atherosclerotic coronary artery plaques. The Tampere Sudden Death Study

**DOI:** 10.1101/2025.01.15.25320636

**Authors:** K Sundström, PP Mishra, S Tuomisto, T Ceder, S Goebeler, M Martiskainen, V Lampinen, MJT Ojanen, V Hytönen, T Lehtimäki, PJ Karhunen

**Affiliations:** Faculty of Medicine and Health Technology Tampere University, Fimlab Laboratories Ltd., Wellbeing Services County of Pirkanmaa, and Finnish Cardiovascular Research Centre Tampere, Tampere Finland; Tampere Genomics Facility, Tampere University, Tampere, Finland; Finnish Institute for Health and Welfare, Tampere, Finland; Laboratory of Protein Dynamics Faculty of Medicine and Health Technology Tampere University, Fimlab Laboratories Ltd. Wellbeing Services County of Pirkanmaa, Tampere, Finland

## Abstract

**Background:** Molecular microbial techniques have identified several DNA sequences from oral and gut bacteria in atherosclerotic plaques. The composition of the plaque microbiome has shown great heterogeneity due to varying DNA extraction and amplification methods, small size sample cohorts, and a low abundance of bacteria in plaques, leading to interference by contaminating bacterial sequences from laboratory chemicals. We used a novel next-generation sequencing (NGS) approach to analyze the entirety of bacterial DNAs found in coronary artery plaques in a unique prospective autopsy series of out-of-hospital deaths representing a cross-section of the general population.

**Methods:** DNA was extracted from aseptically collected and frozen (−80 ^°^C) atherosclerotic coronary plaque samples taken from the left anterior descending (LAD) coronary arteries of 202 victims of sudden out-of-hospital death included in the Tampere Sudden Death Study. Bacterial DNA was amplified using nested 16S rRNA PCR and sequenced. Amplicon sequence variants (ASVs) were inferred using the DADA2 approach. Taxonomy assignment of the sequences was accomplished using the Silva database (version 138).The presence of bacteria in coronary artery atherosclerotic plaques was confirmed by immunohistochemistry.

**Results:** ASVs from 230 bacteria at the genus level occurring in humans were detected in the coronary plaques. The most common ASVs detected in almost all plaques belonged to the oral biofilm-producing bacteria *Veillonella* and *Streptococcus*, which, along with *Prevotella, Lactobacillus, Clostridium*, and *Fusobacterium* were the genera with the most ASV reads following nested PCR. The immunopositivity of oral streptococci and *Veillonella* occurred mainly as biofilm-like structures around calcific plaque areas and associated with the severity of atherosclerosis.

**Conclusion:** Coronary plaques harbor DNA sequences from dozens of mainly oral bacteria. However, only a few bacteria are so common that they likely have a role in the buildup of the microbiome inside a coronary artery atheroma.

## Introduction

Genome-wide association analyses have confirmed that individual susceptibility to coronary heart disease is linked with lipid metabolism and inflammation^1^. The current paradigm suggests that oxidatively modified low-density lipoprotein (LDL) is the primary driver of inflammation^2,3^. The possible role of bacterial infection, although previously rejected due to the failure in large antibiotic trials^4^, has recently gained more interest, as the presence of bacterial DNA fragments in atherosclerotic plaques has been demonstrated with the aid of targeted microbial techniques^5–9^.

Using broad-range 16S rDNA PCR, followed by cloning and sequencing, our research group was the first to report in 2005 that coronary atheromas collected at autopsy contained DNA of bacteria commonly found in the oral cavity^5^, as has now been confirmed by several other studies^6–8^. The recent approach using 16S rRNA gene sequencing–based metagenomic studies has revealed that atherosclerotic plaques harbor DNA sequences from dozens or even hundreds of oral, intestinal, or environmental bacterial genera^8,10,11^, suggesting the presence of a microbiome. However, there is no certainty as to the composition of coronary bacterial plaque microbiota, due to the bias caused by the need to pre-select the bacteria of interest for qPCR, various NGS library preparations and bioinformatics approaches, as well as small cohort sizes. Additionally, the low abundance of bacterial sequences in plaques, can cause amplification of bacterial residues from PCR reagents and other laboratory equipment and environmental elements without clinical significance^12,13^. Therefore, a single round of PCR may not be enough to achieve a reliable microbial profile^13^.

The aim of this study is to apply a novel NGS method using nested PCR to reveal the entirety of the bacterial community in a large population-based cohort of healthy and atherosclerotic coronary artery plaque samples. To confirm the NGS results, we performed bacterial immunohistochemical staining of the coronary plaques.

## Materials and methods

### Study series

We studied the bacterial DNA composition in atherosclerotic plaques in 202 coronary artery plaque samples taken from the left anterior descending arteries of the Tampere Sudden Death Study (TSDS) cohort. The TSDS comprises a prospective series of autopsies on men and women aged > 35 years, who died out of hospital in Tampere and were subjected to a medicolegal autopsy at the Department of Forensic Medicine, Faculty of Medicine and Health Technology, Tampere University. The Tampere region has a population of approximately 0.3 million. None of the cases had died of an infection. In Finland, a complete medicolegal autopsy is performed in all cases of an unexpected out-of-hospital death of a person with no history of a serious disease, or if the person may have died of unnatural causes. The samples were processed at the bacterial laboratory of the Department of Clinical Chemistry, Tampere University. The TSDS protocol was approved by the Ethics Committee of the Pirkanmaa Hospital District (permission number R09097) and the National Supervisory Authority for Welfare and Health.

### Coronary plaque samples

The pericardium was opened aseptically using NaOH + H_2_O + EtOH–treated instruments and sterile gloves. The coronary arteries were opened and dissected free from the surface of the heart on a sterile surgical covering using sterile instruments and stored on sterile Petri dishes, on ice and under cover, until transfer to the laboratory during the same day. In the laboratory, after photographing and measuring the surface areas of atherosclerosis, transverse sections from the most severe and/or thrombotic coronary plaque from the left anterior descending coronary artery (LAD) and right coronary artery (RCA) were collected for DNA analyses and histology under aseptic conditions and with sterile instruments. Tissue samples for DNA were frozen and kept in -80^°^C until analysis. Histological samples were fixed in 10%buffered sterile formalin for 24 hours and then incubated in sterile EDTA solution for 1–2 weeks to decalcify the coronary plaques. Decalcified coronary samples were processed onto slides, stained with the Verhoeff HE method, and classified according to the 1995 terminology of the American Heart Association (AHA) ^14,15^

### DNA extraction and sequencing

DNA was extracted from the frozen LAD coronary samples with a Macherey-Nagel Nucleospin Tissue kit (Macherey-Nagel Gmbh & Company KG, Germany). The coronary samples were chosen based on AHA types in order to have cases with both early- and late-stage atherosclerosis for analysis. DNA extraction was performed according to kit specifications with an extra lysis step, where NextAdvance stainless steel beads (0.9–2.0 mm blend, RNAse-free) were added to the lysis solution and the samples were homogenized with Bullet Blender (NextAdvance, USA) for 6 minutes, level 10, until the tissue was completely homogenized.

Library preparation of the TSDS coronary plaque samples was executed by amplifying the bacterial DNA with nested 16S rRNA PCR and dilution before a second PCR round, according to Sillanpää et al.^16^ with modification. The amplification product was purified with Agenocourt Ampure XP magnetic beads (Life Technologies), and index-PCR was run according to the 16S Metagenomic Sequencing Library Preparation protocol by Illumina^21^ with a Nextera XT Index kit V2 (Illumina Inc.). The amplification products were visualized on 2%agarose gel, and the quality of fragments was assessed with Fragment Analyzer (Advanced Analytical Technologies). Size-selected pools were adjusted to 4 nM using a Qubit 3.0 fluorometer (Life Technologies) and loaded on an Illumina Miseq Reagent kit v2 (500 cycles) (Illumina Inc.) at 4 pM.

### Read data processing and analysis

The quality of the sequence data was inspected using FastQC ^17^ and MultiQC ^18^. Primers from the sequence reads were removed using Cutadapt ^19^. The sequence reads were denoised and dereplicated using functionalities implemented in the DADA2 R/Bioconductor package with the default setting ^20^. Similarly, putative chimeric sequences were also filtered out using DADA2. Three hundred and forty-six genus-level amplicon sequence variants (ASVs) were inferred from the remaining sequences using the DADA2 approach. The taxonomy assignment of the sequences was carried out using the SILVA reference database (version 128) ^21^. Bacterial contaminants from DNA extraction kits and other laboratory reagents were identified and removed manually using a priori knowledge ^12^. In order to calculate the frequencies of ASVs among the 202 TSDS cases, the ASV count data were transformed into proportions using functions implemented in the phyloseq R/Bioconductor package ^22^. The ASV proportion matrix was then converted into a binary matrix by regarding the ASVs found at a proportion greater than zero as present (marked as 1) and others as absent (marked as 0). Then, for every ASV, the frequency was calculated by taking the sum of the ASVs across the TSDS cases in the binary matrix and dividing it by the total number of TSDS samples (n = 202). We also calculated the proportions of each of the ASVs among the detected microbial community in the 202 TSDS cases. For this purpose, we pooled the ASVs across all TSDS cases and transformed their overall abundance into proportions. These ASV proportions represent their abundance in proportion to the microbial community observed among all of the TSDS cases. For the major genus-level ASVs (those with frequency > 50%, n = 18), we also analyzed their relative abundance among the 202 TSDS cases.

### Immunohistochemistry

To confirm the NGS results, we performed bacterial immunohistochemical staining of the coronary plaques with antibodies against the most common bacterial genera observed, following exclusion of contaminating bacterial species originating from DNA extraction kits and other laboratory reagents. The antibodies were produced by ThermoFisher using a 90-day rabbit immunization protocol. The performance and specificity of the resulting antibodies was confirmed with Western blot by staining the respective bacterial lysates (data not shown). We tested the specificity of the antibodies to detect bacterial components in tissue samples according to our previous study ^23^ using rabbit IgG isotype (ABCAM) control, which is a primary antibody that lacks specificity to the target, as well as a rabbit *E. coli*–specific antibody (ABCAM), which both should be negative.

## Results

Of the 202 autopsy cases included in the Tampere Sudden Death Study (TSDS) series, 158 (78.2%) were men and 44 (21.8%) women, with a mean age of 63.6 years (**Table 1**). Most victims (80.2%) had died of a disease, with coronary heart disease being the most common culprit.

**Table 1.** Characteristics of the Tampere Sudden Death Study series (n = 202)

|  |  |
| --- | --- |
| Age | 63.6 (SD 15.6) yrs |
| Sex M/F | 158/44 |
| Body Mass Index (BMI) | 29.2 (SD 7.0) kg/m<sup>2</sup> |
| Postmortem Interval | 4.5 (SD 1.9) days |
| Hypertension | 73 (36.1%) |
| Diabetes | 48 (23.8%) |
| Smoking | 72 (35.6 %) |
| Cause of death |  |
| Disease | 162 (80.2%) |
| Accidental | 16 (7.9%) |
| Suicide | 24 (11.9%) |
| Coronary atherosclerosis (AHA types) <sup>1,2</sup> |  |
| I-III | 49 (24.2%) |
| IV | 30 (14.9%) |
| V | 98 (48.5%) |
| VI | 25 (12.4%) |

According to AHA lesion types, coronary atherosclerosis varied from type I (normal) through V (fibroatheroma with or without calcification) up to VI (atheroma with hemorrhage or thrombosis).

### Bacterial amplicon sequence variants (ASVs)

The total number of bacterial ASVs detected in the coronary atheromas was 346. However, after the manual removal of environmental bacterial contaminants most probably originating from DNA extraction kits and other laboratory reagents, such as Aquabacterium and Pedobacter, as well as combining different variants of the same genus (e.g., *Prevotella*_6 and *Prevotella*_7) into one genus, ASVs from a total of 230 human bacteria at the genus level were detected in the coronary plaques. The 50 most frequent genera are shown in **Figure 1**.

**Figure 1.**
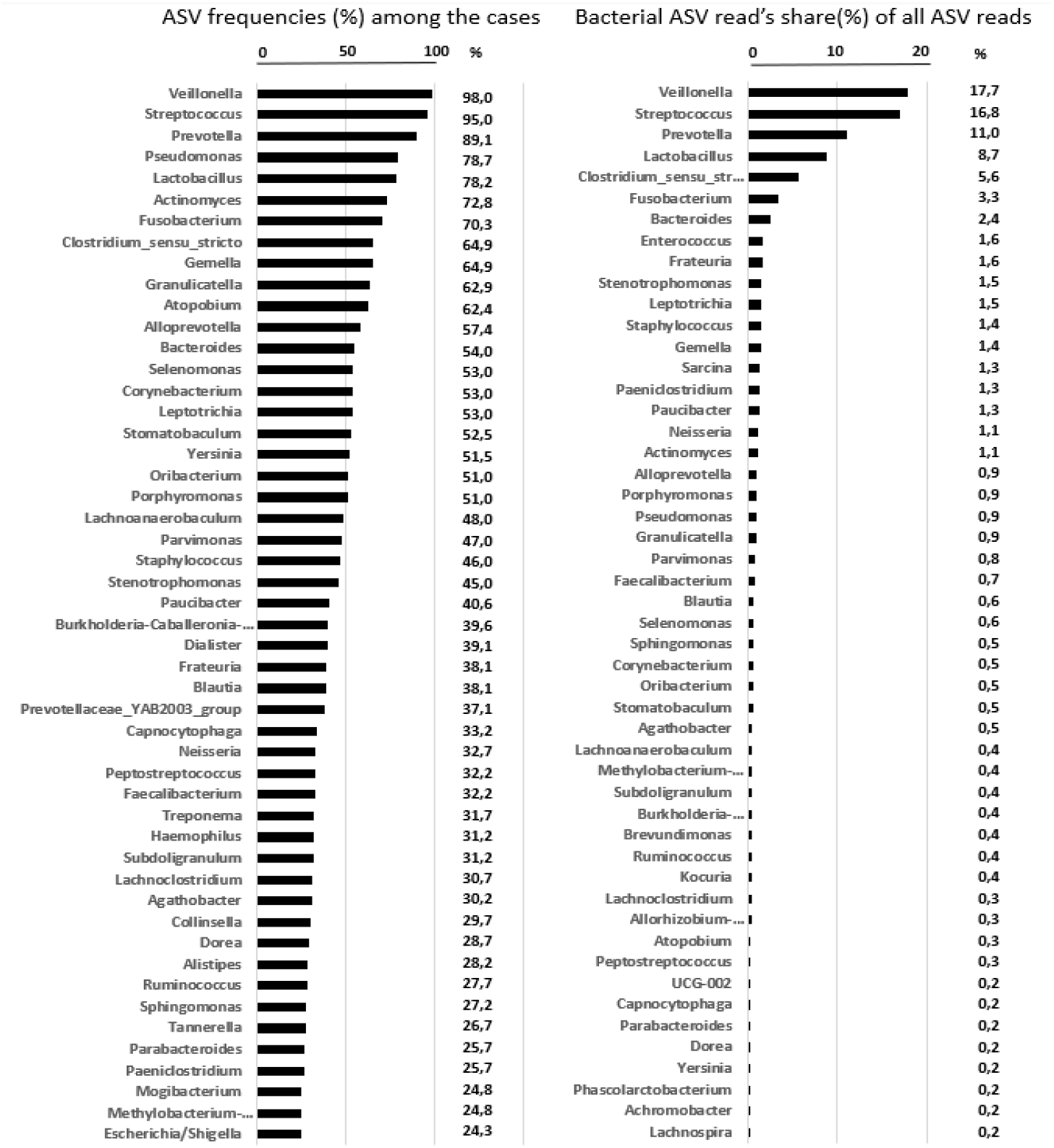
Frequency of amplicon sequencing variants (ASVs) among the coronary atheroma samples of the 202 Tampere Sudden Death victims (left) and the proportions of ASV reads for different bacterial genera out of all ASV reads (right) among the 50 most frequent bacterial genera identified.

ASVs from 20 bacterial genera were present in more than 50%of the plaques. The most common ASVs identified in almost all coronary atheromas belonged to oral biofilm-forming bacteria, such as *Veillonella* (98.0%), followed by *Streptococcus* (95.0%), *Prevotella* (89.1%), *Lactobacillus* (78.2%), *Actinomyces* (72.8%), and *Fusobacterium* (70.3%). DNA sequences from typical gut bacteria that are more seldom found in the oral cavity, such as *Pseudomonas* (78.7%) and *Clostridium* (64.9%), were also detected. DNA from the *Porphyromonas* genus was present in 51.0%of the plaque samples.

Following nested PCR, only 6 bacterial genera commonly found in the oral cavity accounted for more than half (63.1%) of the total number of ASVs. The most frequent ASVs belonged to the *Veillonella* genus (17.7%), followed by *Streptococcus* (16.8%), *Prevotella* (11.0%), *Lactobacillus* (8.7%), *Clostridium* (5.6%), and *Fusobacterium* (3.3%). The remaining ASVs were divided among approximately 200 genera mostly comprising less than 1%of the sequence variant reads. For instance, the proportion of ASVs identified as *Porphyromonas* was 0.7%.

The dominating role of *Veillonella* and *Streptococcus* among the most frequent bacterial genera in the microbiome of individual coronary plaques is seen clearly when the cases are sorted according to increasing read percentage of *Veillonella* ASVs (**Figure 2**).

**Figure 2.**
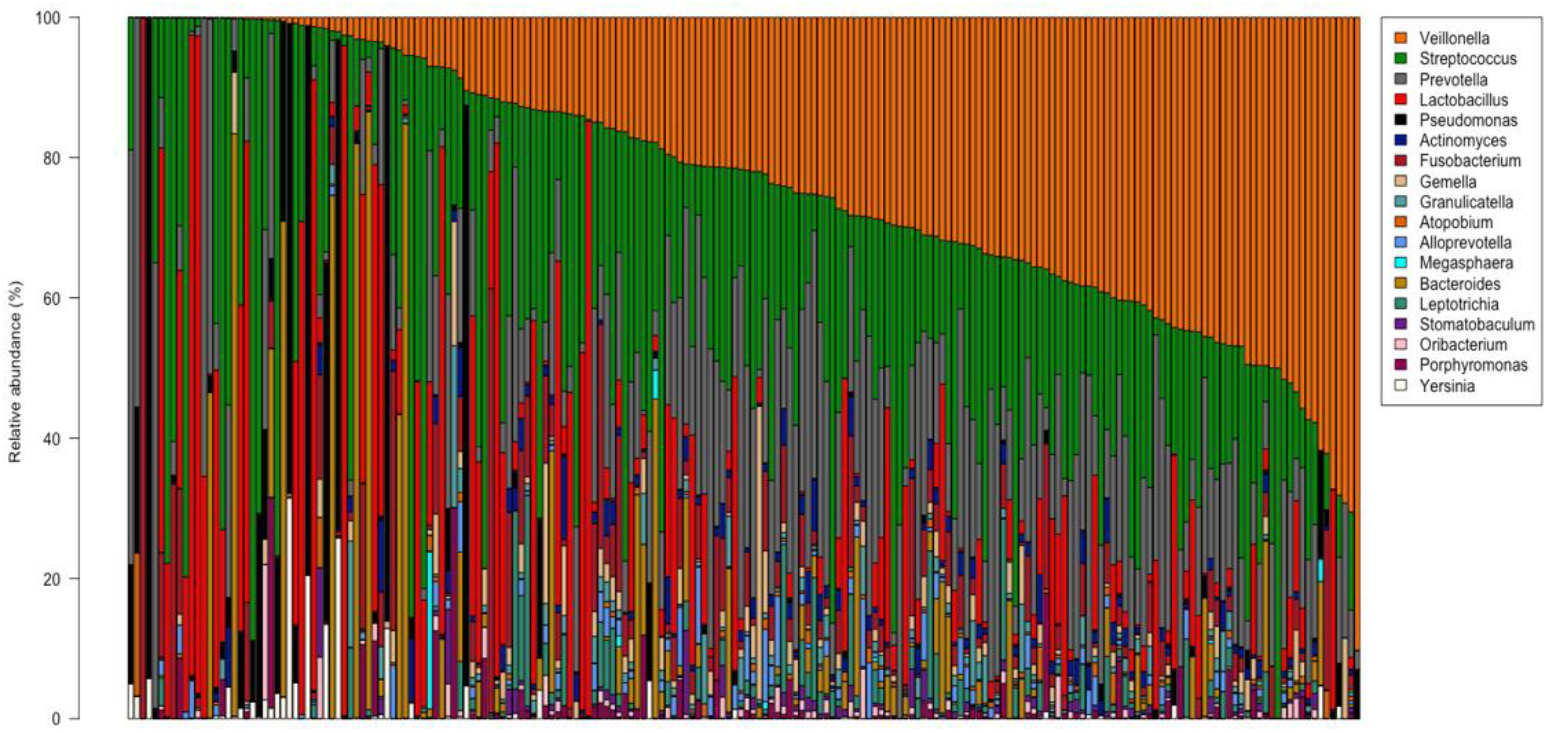
Relative abundance of the 18 most important bacterial taxa at the genus level occurring in > 50%of the coronary plaques of the Tampere Sudden Death Study series (n = 202), grouped according to increasing percentage of amplicon sequence variant (ASV) reads for *Veillonella*.

### Bacterial immunohistochemical staining

Based on the NGS findings of the present study, as well as on our earlier studies ^5,23–27^, we raised polyclonal rabbit antibodies against three major viridans streptococcal species—*Streptococcus sanguinis* (American Type Culture Collection [ATCC] 10556), *Streptococcus mitis* (ATCC 49456), and *Streptococcus gordonii* (ATCC 10558)—as well as *Veillonella parvula* (ATCC 17742), which is the most common species in supragingival and subgingival plaque samples ^28^. We immunostained formalin-fixed and paraffin-embedded TSDS coronary plaques with the antibody cocktail of the three viridans streptococci, as well as with the *Veillonella parvula* antibody, using standard immunohistochemical procedures. Of the coronary plaques stained with the viridans streptococcal cocktail (n = 196), 166 (84.7%) were histologically immunopositive, and of the plaques stained with the *Veillonella* antibody (n = 187), 153 (81.8%) were immunopositive.

### Association of the bacterial immunohistochemical score with coronary plaque atherosclerosis

The immunostaining intensity was assessed with a semiquantitative score (0/+/++/+++) ^23^. There was a strong association (**Figure 3**) between coronary plaques showing advanced atherosclerosis classified as AHA type V (fibrotic or calcified atheroma) or VI (complicated atheroma with rupture and thrombosis/hemorrhage) and the immunopositivity score for viridans streptococci (p < 0.001) and *Veillonella* (p < 0.001). Of the AHA Type I-III coronary samples, 50%of Veillonella-stained and 40%of viridans streptococci–stained plaques were immunopositive. However, of the healthiest coronary samples (AHA Type I), only 10%of Veillonella-stained and 40%of viridans streptococci–stained plaques were immunopositive.

**Figure 3.**
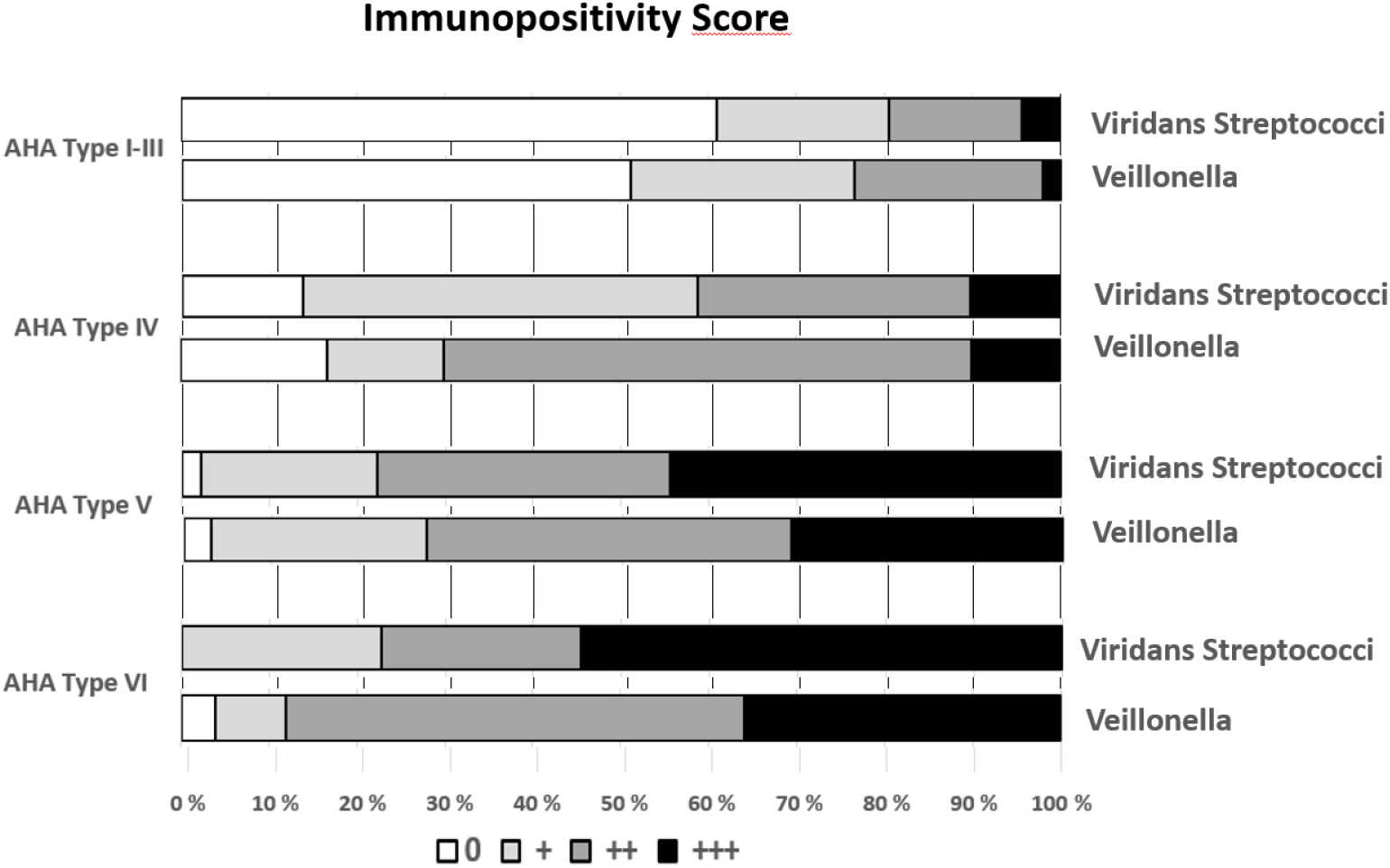
Immunopositivity score of 202 coronary plaques classified by AHA types.

Viridans streptococcal and *Veillonella* immunopositivity seemed to locate as overlapping biofilm-like structures bordering the calcified plaque areas (AHA Type V) in the wall of the stenosed coronary artery (**Figure 4**). ImageJ software (github.com/fiji/fiji) was used to create color convolution micrographs in order to increase the contrast in the immunostained samples for a better visualization of the results ^23^.

**Figure 4.**
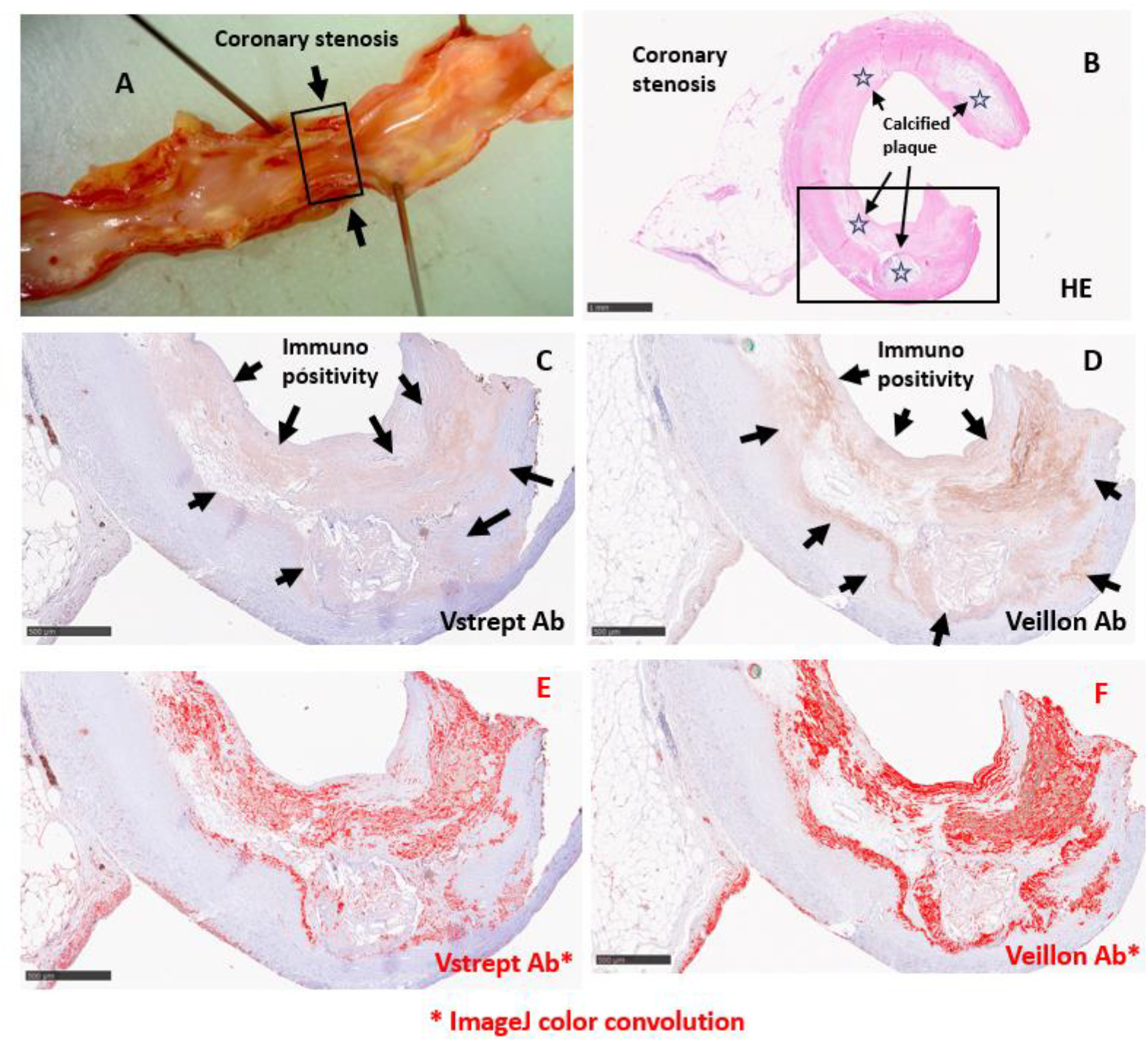
Stenosis of the left anterior descending (LAD) coronary artery (A-B) of a 60-year-old male who died due to coronary artery disease with stenosed and calcified (☆) coronaries and old myocardial infarction. Staining for viridans group streptococci (C) as well as for Veillonella (D) was strongly positive. In color convoluted (Fiji-ImageJ) pictures (E and F) immunopositivity seemed to locate as partly overlapping, biofilm-like structures bordering the calcifying atheromatous lesions.

No streptococci or *Veillonella* immunopositivity was seen in the healthy coronary section taken 3 cm distally from the calcified stenosis site of the same coronary (**Figure. 5**). Control staining performed with a rabbit IgG isotype control antibody (ab37415, Abcam) and with a rabbit IgG anti–Escherichia coli antibody (ab137967, Abcam) was also negative.

**Figure 5.**
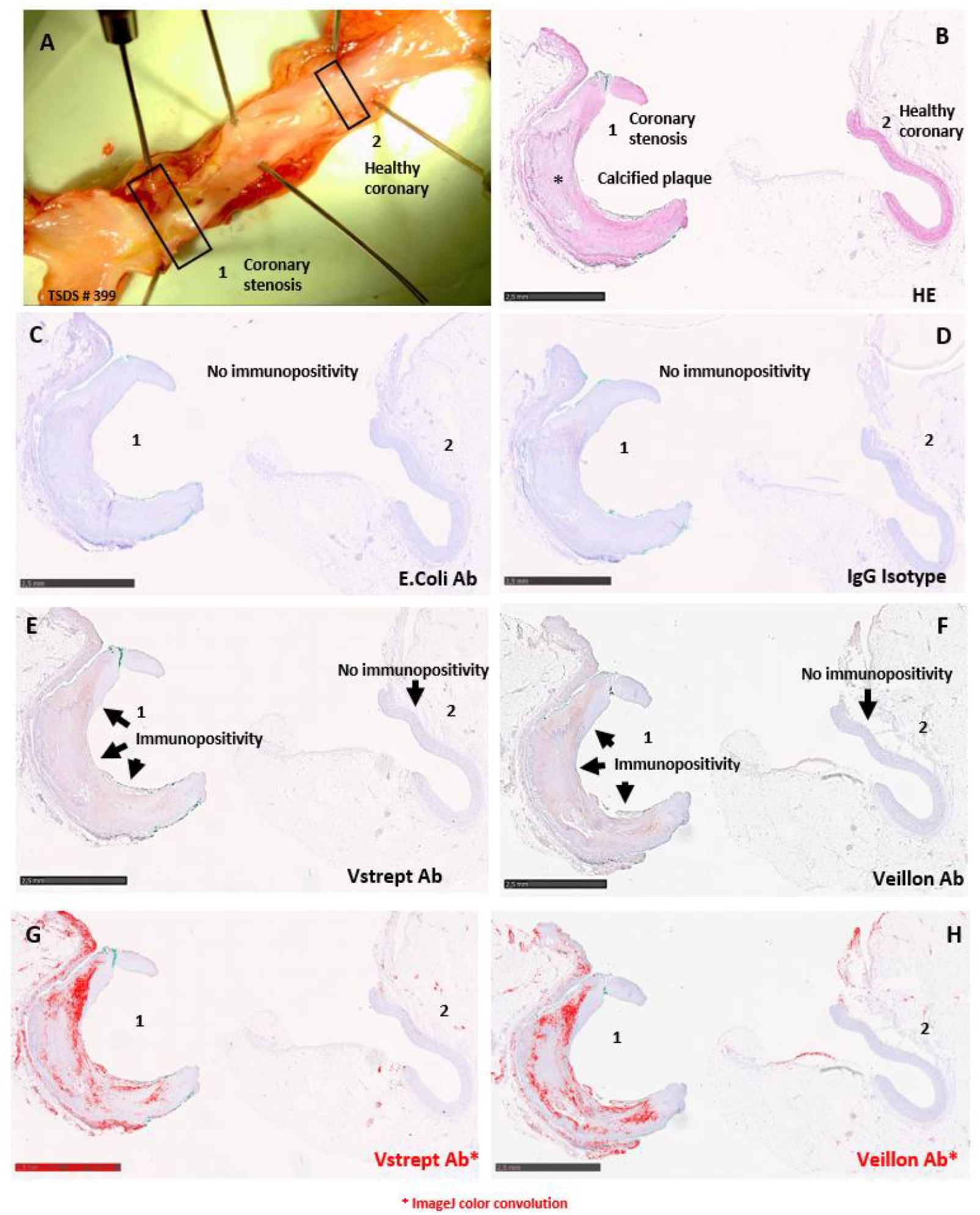
Left coronary artery of a 67-year-old male who died accidentally (A). Coronary samples were taken from the stenosis site (1) as well as from a healthy section (2) 3 cm apart. Hematoxylin - eosin (HE) staining (B) shows AHA type V atherosclerosis with a calcified plaque (**\***) in the stenosis site as well as a healthy coronary section. Control immunohistochemical staining of the calcific plaque and healthy section with an anti-E. Coli antibody (C) as well as with an IgG isotype control antibody (D) were negative. Staining for viridans group streptococci (E) as well as for *Veillonella* (F) was strongly positive, whereas the healthy sections showed no immunopositivity. The immunopositivity is more clearly seen in color convoluted (Fiji-ImageJ) pictures (G-H).

## Discussion

Since the invention of bacterial DNA–based detection techniques, DNA of dozens of different bacteria have been discovered in coronary plaques, suggesting the presence of a bacterial microbiome ^5–11,29,30^. However, the composition of the coronary microbiome has remained uncertain due to sparse sampling and diverse methodology used to detect the bacteria. To generate comprehensive understanding of the bacterial species present in coronary samples, we applied 16s rRNA sequencing, using nested PCR for the library preparation. The use of nested PCR enabled us to detect the entirety of all bacterial DNA sequences from a representative series of 202 coronary plaques obtained during autopsies on victims of sudden out-of-hospital death. After eliminating environmental bacteria that were most probably derived from the PCR reagents, we identified in the coronary plaques a total of 230 bacterial DNA fragments belonging to bacteria occurring in humans at the genus level. However, after nested PCR, 6 bacterial genera had a combined proportion of more than half of the total number of ASV reads, suggesting high abundance and clinical significance. The remaining ASVs consisted of large number of species with small proportion per species (<1%each). The use of nested PCR in our study may have effectively multiplied the more frequent bacterial genera, thus exaggerating the differences in the real ratios of the bacterial DNA fragments in coronary plaques ^31^. However, the advantage of nested PCR in samples with a low bacterial content is supported by our success in detecting bacterial DNA fragments in all analyzed samples, whereas other studies in which nested PCR was not employed have failed to find enough bacterial DNA in most cases ^32,33^. We were able to confirm the presence of bacteria in the coronary samples, using immunohistochemistry with antibodies raised against whole bacteria in rabbits. One further possible explanation for the low number of reads for the most of the bacteria is the possibility that sensitive nested PCR protocol amplifies fragments of bacterial DNA from monocytes and macrophages present in the coronary atheroma at the time of death, carrying phagocytized bacterial DNA originating from a noncardiac site, such as the gingiva, the skin, or the respiratory tract ^34^.

The most frequent ASVs in our study belonged to the *Veillonella* genus, followed by *Streptococcus, Prevotella, Lactobacillus, Clostridium*, and *Fusobacterium*. These bacteria are normal inhabitants of human saliva ^35^ and participate in the formation of the mixed-species communities attaching sequentially to the surface of the teeth ^36,37^, forming early, middle, and late dental plaque, ultimately leading to the development of dental calculus ^38^. Streptococci, - the first colonizers - produce significant amounts of lactic acid, and veillonellae utilize lactic acid as their source of carbon and energy. This metabolic connection has been suggested to be a driving force in the development of multispecies communities and in the transition of initial colonizers as promoters of early and middle colonization ^39^. While *Streptococcus, Veillonella*, and *Prevotella* are known to act as early colonizers in a dental biofilm, *Lactobacillus* and *Clostridium* belong to the oral microbiome in adults with dental caries ^40^. Clostridium is also one of the most represented anaerobic bacteria in healthy gut ^41^. DNA sequences of *Pseudomonas* were found in almost all our coronary atheromas but covered only 0.9%of the total ASV reads. *Pseudomonas* has been suggested as a possible reagent contaminant ^12^ but it has been found in the gut and also in the mouth of middle-aged patients, showing no relation to inflammatory processes in the oral cavity ^42,43^. The *Bacteroides* genus was present in 54%of the plaques, and its ASV reads comprised 2.4%of the total proportion of the ASV reads. In the oral cavity, a certain *Bacteroides* genus has been associated with gingivitis, periodontitis, endodontal infections, and odontogenic abscesses^44^. The relative abundance of different oral bacterial taxa in our coronary plaques closely resembled the distribution of the same bacterial taxa in normal human saliva^35^. Koren et al.^7^ analyzed 15 carotid endarterectomy samples with 454 pyrosequencing and, in line with our results, found P*seudomonas* in all atherosclerotic plaque samples, and *Veillonella* and *Streptococcus* in the majority of them. Interestingly, in their study, the combined abundances of *Veillonella* and *Streptococcus* in atherosclerotic plaques correlated with their abundance in the oral cavity. They did not find *Chlamydia* by means of pyrosequencing, nor did we detect any ASVs for *Chlamydia* in our series, suggesting its minor significance. All of these findings indicate that the bacterial microbiome in coronary plaques closely resembles the oral microbiome and consists of bacteria known to be involved in biofilm formation.

The use of sequencing-based metagenomic analysis methods has the great advantage that all bacterial DNA in the target tissue is analyzed and the clinical hypothesis or interests of the study group do not bias the results, as might be the case with qPCR-based methods where the primers target only specific bacteria. This methodological selection bias may be the reason why the main bacteria of interest so far have been *Porphyromonas gingivalis, Chlamydia pneumoniae*, and *Helicobacter pylori* ^45^. Moreover, most arterial plaque analyses using targeted primers are from very small cohorts. For example, Nakano et al.^29^ detected *Streptococcus mutans* with broad-range PCR in 74%of the 27 studied plaques, which were collected from thoracic or abdominal aortic aneurysms. Rath et al.^30^ identified *Aggregatibacter actinomycetemcomitans* in five out of seven endarterectomy plaque samples with PCR using specific primers. Most arterial plaque bacteria analysis samples are derived from endarterectomy ^7,8,11,46^, which is performed only for patients suffering from symptomatic disease. Our samples consist of a cross-section of the entire left ascending coronary artery tissue, whereas endarterectomy samples are restricted to the intima and removed plaque tissue. Our study cohort is also considerably larger compared to other studies ^7,8,10,11^, ranging from healthy tissues to very calcified and even ruptured plaques. We sequenced the samples with massive parallel sequencing technology, the Illumina’s MiSeq sequencer, instead of older sequencing technologies, such as the pyrosequencing ^7,11^, qPCR, or DGGE ^6^ used in previous studies of atherosclerotic plaque microbiota. We eliminated all environmental bacteria, such as the common *Aquabacterium*, from our analysis. Furthermore, the most common genera did not include staphylococci, for example, indicating that there was no cross-contamination from the skin of the laboratory personnel.

Immunohistochemical studies using antibodies against three species of viridans-group streptococci and *Veillonella parvula* confirmed our bacterial DNA findings. The immunopositive areas formed biofilm-like structures. The immunopositivity score was strongly associated with calcified coronary plaques (AHA type V) and complicated coronary plaques showing thrombosis or intraplaque hemorrhage (AHA type VI), suggesting that the plaque microbiome may be involved in the pathogenesis of a myocardial infarction.

The strengths of the study include the unique population-based autopsy series of human coronary arteries with histologically defined atherosclerotic plaque severity (AHA classification), as well as the considerably larger sample size compared to earlier studies, the nested PCR–based NGS metagenomic method which takes into account a wider array of bacteria than earlier techniques, and the careful consideration and filtering of the possible contaminating environmental bacteria. The strength is also the fact that we could confirm the presence of most the common viridans streptococci and Veillonella bacterial DNA findings by immunohistochemistry in same coronary sections using antibodies raised against these bacteria. The limitation is that the TSDS cohorts included in this study is of European origin, and, therefore, studies with populations of different ethnicities are needed. The possibility of postmortem contamination or bacterial contamination during storage of the samples seems unlikely because in that case also the healthy coronary segments within the same histological block should carry immunohistochemically detectable bacteria.

In conclusion, coronary artery plaques harbor DNA sequences from dozens of mainly oral but also gut bacteria. However, only a few oral bacteria constitute most of the reads, suggesting that they may have a key role in the buildup of the microbiome and biofilm-like structures inside a coronary atheroma. Those species appear as attractive targets for preventive measures.

## Data Availability

All data will be available upon reasonable request

## Acknowledgements

The Tampere Sudden Death Study (TSDS) was financially supported by EU’s 7th Framework Programme (grant no. 201668 for AtheroRemo); the Jane and Aatos Erkko Foundation; the Tampere University Foundation; the Tampere University Hospital Medical Funds; the Finnish Foundation of Cardiovascular Research (K.S., P.K.,T.L. and V.H.); the Pirkanmaa Regional Fund of the Finnish Cultural Foundation (K.S.); the Yrjö Jahnsson Foundation; the Juho Vainio Foundation; the Sigrid Juselius Foundation (V.H.) the Tampere Tuberculosis Foundation; EU Horizon 2020 (grant 755320 for TAXINOMISIS and grant 848146 for To Aition); the Academy of Finland (grants 322098 and 356405 for T.L., 331946 for V.H.); the Tampere University Hospital Supporting Foundation; and the Finnish Society of Clinical Chemistry. Pashupati P. Mishra was supported by the Academy of Finland (grant number 349708). We acknowledge Biocenter Finland for infrastructure support.

## References

1. Consortium, Cardi. et al. Large-scale association analysis identifies new risk loci for coronary artery disease. Nature genetics 45, 25–33 (2013).

2. Binder, C. J. et al. Innate and acquired immunity in atherogenesis. Nature medicine 8, 1218–1226 (2002).

3. Libby, P., Ridker, P. M. & Hansson, G. K. Progress and challenges in translating the biology of atherosclerosis. Nature 473, 317–325 (2011).

4. Song, Z., Brassard, P. & Brophy, J. M. A meta-analysis of antibiotic use for the secondary prevention of cardiovascular diseases. The Canadian journal of cardiology 24, 391–395 (2008).

5. Lehtiniemi, J., Karhunen, P. J., Goebeler, S., Nikkari, S. & Nikkari, S. T. Identification of different bacterial DNAs in human coronary arteries. European journal of clinical investigation 35, 13–16 (2005).

6. Ott, S. J. et al. Detection of diverse bacterial signatures in atherosclerotic lesions of patients with coronary heart disease. Circulation 113, 929–937 (2006).

7. Koren, O. et al. Human oral, gut, and plaque microbiota in patients with atherosclerosis. Proceedings of the National Academy of Sciences of the United States of America 108 Suppl, 4592–4598 (2011).

8. Mitra, S. et al. In silico analyses of metagenomes from human atherosclerotic plaque samples. Microbiome 3, 38-015-0100-y (2015).

9. Tuomisto, S. et al. Age-dependent association of gut bacteria with coronary atherosclerosis: Tampere Sudden Death Study. PloS one 14, e0221345 (2019).

10. Mougeot, J. C. et al. Porphyromonas gingivalis is the most abundant species detected in coronary and femoral arteries. Journal of oral microbiology 9, 1281562 (2017).

11. Ziganshina, E. E. et al. Bacterial Communities Associated with Atherosclerotic Plaques from Russian Individuals with Atherosclerosis. PloS one 11, e0164836 (2016).

12. Salter, S. J. et al. Reagent and laboratory contamination can critically impact sequence-based microbiome analyses. BMC biology 12, 87-014-0087-z (2014).

13. Brun, A. et al. Innovative application of nested PCR for detection of Porphyromonas gingivalis in human highly calcified atherothrombotic plaques. Journal of Oral Microbiology 12, 1742523 (2020).

14. Stary, H. C. et al. A definition of advanced types of atherosclerotic lesions and a histological classification of atherosclerosis. A report from the Committee on Vascular Lesions of the Council on Arteriosclerosis, American Heart Association. Circulation 92, 1355–1374 (1995).

15. Virmani, R., Kolodgie, F. D., Burke, A. P., Farb, A. & Schwartz, S. M. Lessons from sudden coronary death: a comprehensive morphological classification scheme for atherosclerotic lesions. Arteriosclerosis, Thrombosis, and Vascular Biology 20, 1262–1275 (2000).

16. Sillanpää, S. et al. Next-Generation Sequencing Combined with Specific PCR Assays To Determine the Bacterial 16S rRNA Gene Profiles of Middle Ear Fluid Collected from Children with Acute Otitis Media. mSphere 2, (2017).

17. FastQC (https://www.bioinformatics.babraham.ac.uk/projects/fastqc/). (2015).

18. Ewels, P., Magnusson, M., Lundin, S. & Käller, M. MultiQC: summarize analysis results for multiple tools and samples in a single report. Bioinformatics 32, 3047–3048 (2016).

19. Martin, M. Cutadapt removes adapter sequences from high-throughput sequencing reads. EMBnet.journal 17, 10–12 (2011).

20. Callahan, B. J. et al. DADA2: High-resolution sample inference from Illumina amplicon data. Nat Methods 13, 581–583 (2016).

21. Quast, C. et al. The SILVA ribosomal RNA gene database project: improved data processing and web-based tools. Nucleic Acids Research 41, D590–D596 (2013).

22. McMurdie, P. J. & Holmes, S. phyloseq: An R Package for Reproducible Interactive Analysis and Graphics of Microbiome Census Data. PLOS ONE 8, e61217 (2013).

23. Patrakka, O. et al. Thrombus Aspirates From Patients With Acute Ischemic Stroke Are Infiltrated by Viridans Streptococci. J Am Heart Assoc 12, e030639 (2023).

24. Pessi, T. et al. Bacterial signatures in thrombus aspirates of patients with myocardial infarction. Circulation 127, 1219–1228 (2013).

25. Pyysalo, M. J., Pyysalo, L. M., Pessi, T., Karhunen, P. J. & Ohman, J. E. The connection between ruptured cerebral aneurysms and odontogenic bacteria. Journal of neurology, neurosurgery, and psychiatry 84, 1214–1218 (2013).

26. Vakhitov, D. et al. Bacterial signatures in thrombus aspirates of patients with lower limb arterial and venous thrombosis. Journal of vascular surgery 67, 1902–1907 (2018).

27. Patrakka, O. et al. Oral Bacterial Signatures in Cerebral Thrombi of Patients With Acute Ischemic Stroke Treated With Thrombectomy. J Am Heart Assoc 8, e012330 (2019).

28. Giacomini, J. J., Torres-Morales, J., Dewhirst, F. E., Borisy, G. G. & Mark Welch, J. L. Site Specialization of Human Oral Veillonella Species. Microbiol Spectr 11, e04042–22.

29. Nakano, K. et al. Detection of cariogenic Streptococcus mutans in extirpated heart valve and atheromatous plaque specimens. Journal of clinical microbiology 44, 3313–3317 (2006).

30. Rath, S. K., Mukherjee, M., Kaushik, R., Sen, S. & Kumar, M. Periodontal pathogens in atheromatous plaque. Indian journal of pathology & microbiology 57, 259–264 (2014).

31. Green, M. R. & Sambrook, J. Nested Polymerase Chain Reaction (PCR). Cold Spring Harb Protoc 2019, (2019).

32. Wang, X. et al. Detecting prokaryote-specific gene and other bacterial signatures in thrombi from patients with acute ischemic stroke. Thromb J 22, 14 (2024).

33. Sato, A. et al. Metagenomic Analysis of Bacterial Microflora in Dental and Atherosclerotic Plaques of Patients With Internal Carotid Artery Stenosis. Clin Med Insights Cardiol 18, 11795468231225852 (2024).

34. Katz, J. T. & Shannon, R. P. Bacteria and coronary atheroma: more fingerprints but no smoking gun. Circulation 113, 920–922 (2006).

35. Sundström, K. et al. Similarity of salivary microbiome in parents and adult children. PeerJ (San Francisco, CA) 8, e8799–e8799 (2020).

36. Kolenbrander, P. E., Palmer Jr, R. J., Periasamy, S. & Jakubovics, N. S. Oral multispecies biofilm development and the key role of cell-cell distance. Nature reviews.Microbiology 8, 471–480 (2010).

37. Periasamy, S. & Kolenbrander, P. E. Mutualistic biofilm communities develop with Porphyromonas gingivalis and initial, early, and late colonizers of enamel. Journal of Bacteriology 191, 6804–6811 (2009).

38. Velsko, I. M. et al. Microbial differences between dental plaque and historic dental calculus are related to oral biofilm maturation stage. Microbiome 7, 102–102 (2019).

39. Periasamy, S. & Kolenbrander, P. E. Central role of the early colonizer Veillonella sp. in establishing multispecies biofilm communities with initial, middle, and late colonizers of enamel. J Bacteriol 192, 2965–2972 (2010).

40. Badet, C. & Thebaud, N. B. Ecology of lactobacilli in the oral cavity: a review of literature. The open microbiology journal 2, 38–48 (2008).

41. Alou, M. T. et al. Taxonogenomic description of four new Clostridium species isolated from human gut: ‘Clostridium amazonitimonense’, ‘Clostridium merdae’, ‘Clostridium massilidielmoense’ and ‘Clostridium nigeriense’. New microbes and new infections 21, 128–139 (2018).

42. Botzenhart, K., Puhr, O. F. & Döring, G. Pseudomonas aeruginosa in the oral cavity: occurrence and age distribution of adult germ carriers. Zentralblatt fur Bakteriologie, Mikrobiologie und Hygiene. 1. Abt. Originale B, Hygiene 180, 471- (1985).

43. Wheatley, R. M. et al. Gut to lung translocation and antibiotic mediated selection shape the dynamics of Pseudomonas aeruginosa in an ICU patient. Nat Commun 13, 6523 (2022).

44. van Winkelhoff, A. J., van Steenbergen, T. J. & de Graaff, J. The role of black-pigmented Bacteroides in human oral infections. J Clin Periodontol 15, 145–155 (1988).

45. Rosenfeld, M. E. & Ann Campbell, L. Pathogens and atherosclerosis: Update on the potential contribution of multiple infectious organisms to the pathogenesis of atherosclerosis. Thrombosis and haemostasis 106, 858–867 (2011).

46. Lanter, B. B., Sauer, K. & Davies, D. G. Bacteria present in carotid arterial plaques are found as biofilm deposits which may contribute to enhanced risk of plaque rupture. mBio 5, e01206–14 (2014).

